# Comparing Continuous versus Intermittent-Threshold Drainage Strategies for Spinal Perfusion Pressure Optimization in Patients with Acute Traumatic Spinal Cord Injuries

**DOI:** 10.1101/2023.08.18.23291810

**Authors:** Raj Swaroop Lavadi, Regan Shanahan, David Kojo Hamilton, Thomas James Buell, Nitin Agarwal, Ava Puccio, David Okwudi Okonkwo, Daryl Pinion Fields

**Affiliations:** Department of Neurological Surgery, University of Pittsburgh School of Medicine, Pittsburgh, PA, USA; Department of Neurological Surgery, University of Pittsburgh Medical Center, Pittsburgh, PA, USA

**Keywords:** Lumbar drain, CSF drainage, Spinal cord perfusion pressure, Spinal cord injury, Vasopressor, Mean arterial pressure

## Abstract

**Study Design:** A cross-sectional study.

**Objective:** The primary objective of this study is to compare the efficacy of continuous versus threshold drainage strategies for maintaining spinal cord perfusion pressure (SCPP) in patients with new traumatic spinal cord injuries (SCI).

**Setting:** Level 1 trauma center.

**Methods:** A retrospective study of 19 patients with traumatic SCIs. SCPP was optimized at the discretion of the managing clinician using either vasopressors to increase mean arterial pressure or cerebral spinal fluid (CSF) drainage to decrease intrathecal pressure. Six patients were managed with continuous drainage (CSF drained at regular intervals regardless of SCPP) and 13 had CSF drained only when SCPP fell below 65mmHg (i.e. threshold drainage). Intrathecal pressure, SCPP, mean arterial pressure, and vasopressor utilization were compared using univariate T-test statistical analysis.

**Results:** The cohort included over 1500 time points from 19 patients. While there was no difference in rates of sub-optimal SCPP (< 65mmHg; p = 0.257), patients managed with threshold drainage were more likely to exhibit critically-low SCPP (< 50 mmHg; p = 0.003) despite also having lower average intrathecal pressures (p < 0.001). There were no differences in average SCPP, MAP, or vasopressor utilization between the two groups (p > 0.05).

**Conclusions:** Acute SCI patients managed with continuous CSF drainage were less likely to exhibit critically-low SCPPs, previously shown to be associated with worse clinical recovery. A larger, prospective cohort is needed to validate the impact of CSF drainage strategies on long-term SCI outcomes.

## INTRODUCTION

There are over 17,000 new spinal cord injuries in the United States each year [1]. Management of traumatic spinal cord injury includes surgery to decompress neural elements and stabilized the spine, followed by medical management to minimize secondary insults; including spinal cord hypo-perfusion related cord ischemia [2].

The hemodynamic management of spinal cord injury is evolving to include a focus on spinal cord perfusion pressure (SCPP) defined as the difference between mean arterial pressure and intrathecal pressure (ITP) [3]. Previous work has linked SCPP < 50mmHg with worse outcomes, [4] while SCPP > 65mmHg in the early post injury phase has been associated with improved neurologic recovery. Patients with SCPP < 65mmHg may either receive vasopressors to drive up their mean arterial pressure (MAP) or have cerebrospinal fluid (CSF) removed through a lumbar intrathecal drain. There is no consensus as to which method is more effective but a recent study highlights the difficulty in maintaining high MAPs in SCI patients with autonomic volatility [5].

CSF drainage can be achieved via two distinct strategies; 1) threshold drainage involves CSF removal only when SCPP drops < 65mmHg, and 2) continuous drainage involves removing 5 – 10mL of CSF at regular one-hour intervals regardless of SCPP. We compared the impact of threshold versus continuous CSF drainage on spinal cord perfusion pressure in a single-center cohort of acute spinal cord injury patients.

## METHODS

### Experimental Protocol

All procedures, including lumbar drain placement, CSF drainage, and collection of relevant clinical data from electronic medical records were approved by the University of Pittsburgh Institutional Review Board (STUDY19070184), with necessary consent provided by the patients. Patients presenting to a single level 1 trauma center between 2018–2022 with cervical or thoracic traumatic spinal cord injury severity grade A–C as evaluated by the ASIA impairment scale (AIS) were eligible for inclusion [6]. All patients underwent spinal stabilization surgery with a lumbar drain placed either intraoperatively or postoperatively in the intensive care unit. All patients were managed at the discretion of the lead clinician for a targeted SCPP > 65mmHg. ITP was monitored using a lumbar intrathecal catheter and MAP was measured using an arterial line. Systemic oxygenation was monitored using surface pulse oximetry. SCPP was calculated as MAP minus intrathecal pressure [7]. Patients managed with continuous drainage had 5-10mL of CSF removed from the intrathecal catheter every hour, regardless of SCPP. Alternatively, patients managed with threshold drainage had CSF withdrawn only when SCPP dropped < 65mmHg. In patients with low MAP (< 60mmHg), a vasopressor (most commonly norepinephrine) was utilized to increase MAP and, thereby, SCPP. Choice of vasopressor and the decision to use vasopressor versus CSF drainage was left to the discretion of the managing clinician. All vasopressors were converted to equivalent norepinephrine dosages as follows; 1:10 norepinephrine:phenylephrine [8].

### Demographics

The two groups demonstrated similar gender and age distributions (p > 0.05). Information on spinal cord injury location, clinical course of neurological function, and patient demographics can be found in **Table 1**.

**Table 1.**
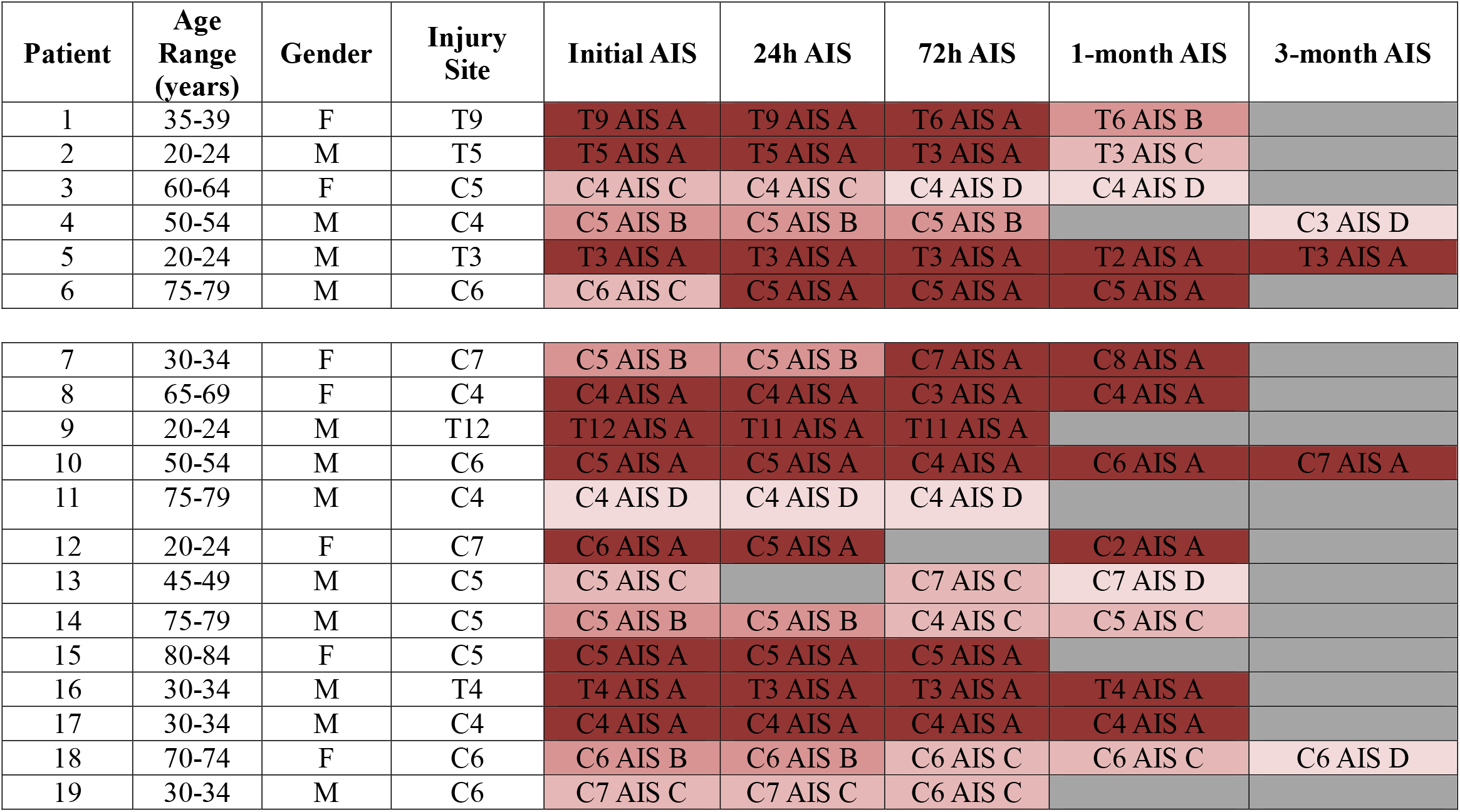
Patient demographics and injury characteristics. Patients 1-6 were managed with continuous drainage and patients 7-19 were managed with threshold drainage. The color gradient represents the transition in AIS scores over time (darker color indicates greater severity). There was no significant difference in patient age between the two groups (p = 0.31). AIS, ASIA (American Spinal Injury Association) Impairment Scale.

### Statistics

Normality was assessed with Shapiro-Wilks test. ITP, SCPP, MAP, and vasopressor utilization were compared using univariate T-test statistical analysis; SAS 9.4 (Cary, NC). Differences were considered significant if p value < 0.05.

## RESULTS

### SCPP

Threshold drainage and continuous drainage groups demonstrated similar average SCPP (p = 0.935), average MAP (p = 0.112), and average systemic oxygenation (p = 0.94). Threshold drainage patients exhibited significantly lower average ITP (p < 0.001; **Table 2**). There was no difference in percentage of time spent below optimal SCPP (< 65mmHg; p = 0.257). Threshold drainage patients were more likely to exhibit critically low SCPPs (< 50mmHg; p = 0.003; **Figure 1**) when compared to continuous drainage patients. There was no difference in vasopressor use between the two groups (p = 0.76; **Figure 2**).

**Table 2.**
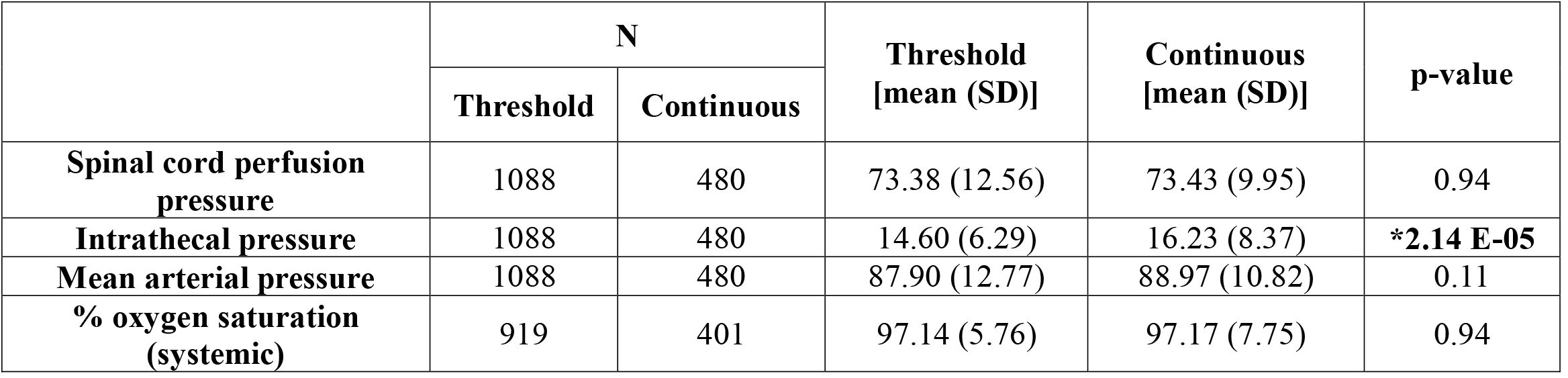
Average subacute pressure and systemic oxygenation. The ‘N’ value represents the number of individual data points collected for each variable. The groups were considered significantly different if p < 0.05 *.

|  | N |  | Threshold<br>[mean (SD)] | Continuous<br>[mean (SD)] | p-value |
| --- | --- | --- | --- | --- | --- |
|  | Threshold | Continuous |  |  |  |
| Spinal cord perfusion pressure | 1088 | 480 | 73.38 (12.56) | 73.43 (9.95) | 0.94 |
| Intrathecal pressure | 1088 | 480 | 14.60 (6.29) | 16.23 (8.37) | **\*2.14 E-05** |
| Mean arterial pressure | 1088 | 480 | 87.90 (12.77) | 88.97 (10.82) | 0.11 |
| % oxygen saturation (systemic) | 919 | 401 | 97.14 (5.76) | 97.17 (7.75) | 0.94 |

**Figure 1.**
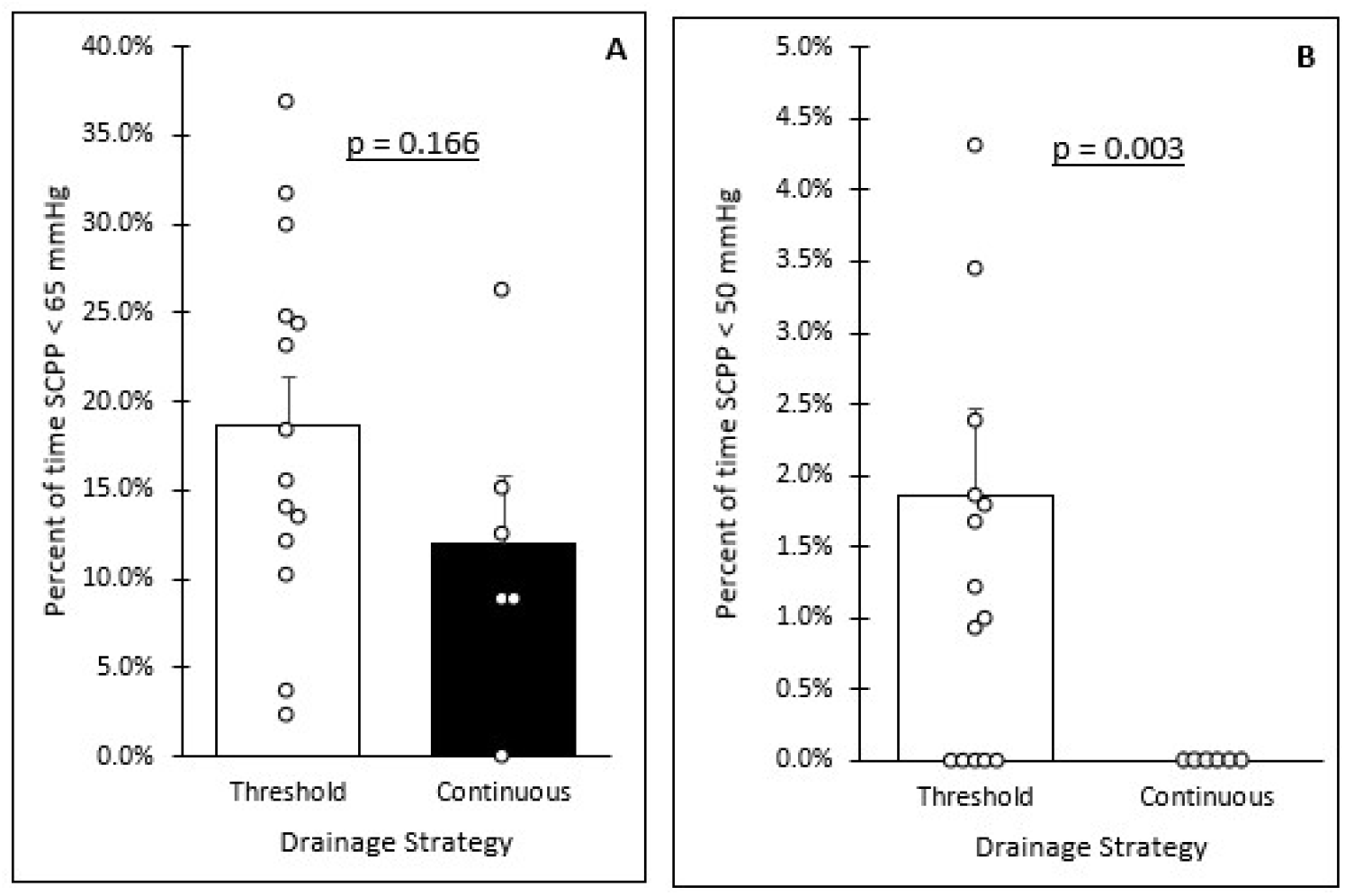
Frequency of suboptimal and critically low spinal cord perfusion pressure. **A)** Comparing percentage of time spent in suboptimal spinal cord perfusion pressure (< 65 mmHg; p = 0. 257). **B)** Comparing percentage of time spent in critically low spinal cord perfusion pressure (< 50 mmHg; p = 0.003). Graphs demonstrate groups averages with error bars representing standard error of mean. The groups were considered significantly different if p < 0.05.

**Figure 2.**
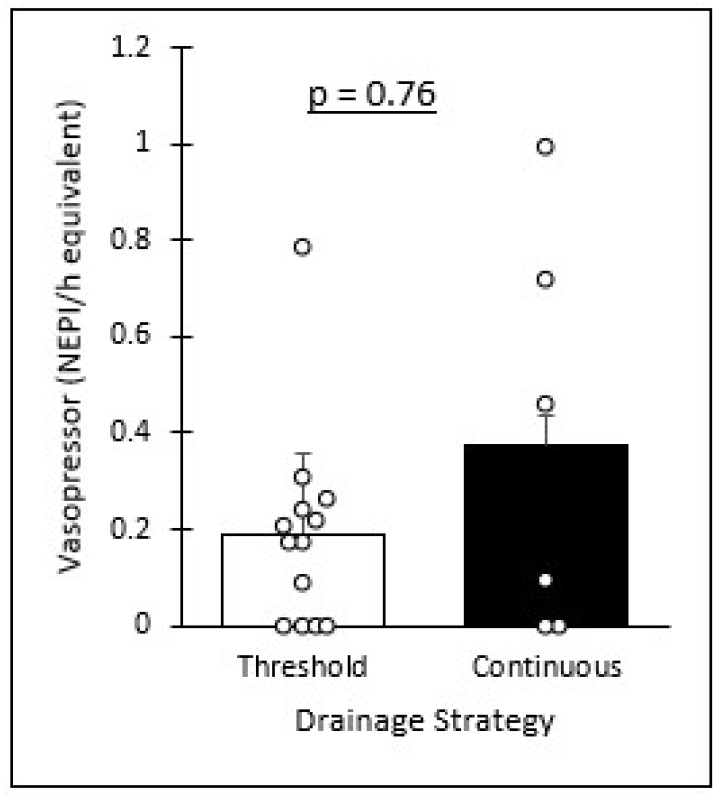
Average vasopressor use. There was no significant difference in vasopressor utilization between the two groups (p = 0.76). Graphs demonstrate groups averages with error bars representing standard error of mean. The groups were considered significantly different if p < 0.05.

### Patient Outcomes

Of the 19 patients included in this study, 15 had documented neurological exams at least one-month after the index injury (six continuous patients and nine threshold patients; **Table 1**). Of the six continuous patients, four demonstrated at least a single grade improvement in their AIS exam, one demonstrated a subacute decline (< 72 hours after injury), and one remained an AIS A at least three-month after injury. Of the 9 patients managed with threshold drainage, three demonstrated at least a single grade improvement in their AIS exam, one worsened from AIS B to AIS A, while the other five patients remained stable with persistent AIS A exams. No patient incurred a complication related to lumbar drain placement, use, or removal.

## DISCUSSION

Within this cohort of 19 spinal cord injury patients, patients managed with continuous drainage were less likely to exhibit critically low SCPP (< 50mmHg). This data highlights the need for further work to refine strategies for secondary insult prevention in spinal cord injury patients.

SCPP represents the pressure difference from mean arterial pressure and intrathecal pressure [7] and is used as a surrogate marker of spinal tissue oxygen delivery. In patients with traumatic spinal cord injuries, critically low SCPPs are associated with worse six-month clinical outcomes. While a causative link has not been confirmed, we theorize that acute hypoperfusion increases the rate of spinal cord infarct that undermines recovery potential. [9] While patients managed with threshold and continuous drainage strategies demonstrate similar average SCPP, and a similar propensity for sub-optimal (< 65mmHg) perfusion pressures, only threshold drainage patients demonstrated critically low SCPPs. The significance of transient (< 1h) spinal hypoperfusion (suboptimal or critical hypoperfusion) is not well-studied and represents a gap in our understanding of secondary insult prevention. Future animal models and human studies must explore spinal tissue tolerance to critical hypoperfusion, and responsiveness to corrective measures.

Prognostic tools that utilize magnetic resonance imaging suggest injury site edema results in worse clinical outcomes [10], likely secondary to local perfusion deficits that result in tissue ischemic injury [11]. The present study does not address cord edema or local tissue perfusion deficits but we can speculate that maintaining appropriate SCPP may indirectly limit local edema-associated ischemic injury. An analogous example would be measures to prevent systemic hypotension in acute stroke patients; global measures to increase perfusion indirectly enhance local perfusion. Interestingly, continuous drainage did not result in a lower intrathecal pressure. This paradoxical observation is beyond the scope of this particular study but warrants further investigation into the link between CSF drainage and intrathecal pressure in spinal cord injury patients.

### Limitations

The small sample size limits our capacity to assess the impact of CSF drainage strategy on neurological outcomes. The small sample size also limits analysis of cervical versus thoracic spinal cord injury as well as injury pattern (fracture/dislocation, central cord syndrome, etc.).

While this study assumes SCPP is representative of spinal cord oxygen delivery, we did not directly test spinal or CSF oxygenation. Placement of an intraspinal oxygen monitor or non-invasive tissue oxygen detector may provide better insight into tissue oxygenation but these tools are not commonly available for clinical use. A similar debate exists in traumatic brain injury. The ongoing Brain Oxygen Optimization in Severe Traumatic Brain Injury Phase-3 (BOOST-3) trial utilizes intracranial-intraparenchymal monitors to guide oxygenation strategies for severe traumatic brain injury patients [12]. The results of the BOOST-3 study will inform us if tissue oxygenation probes are superior to pressure monitors in evaluating oxygen delivery changes, an important consideration for ongoing spinal cord injury trials.

## CONCLUSION

Continuous CSF drainage in traumatic spinal cord injury patients was associated with fewer incidences of critically-low spinal cord perfusion pressure (SCPP < 50mmHg) when compared to threshold CSF drainage. These observations suggest the need for a larger prospective study that explores the long-term clinical impact of alternative CSF drainage strategies in traumatic spinal cord injury.

## Data Availability

All data produced in the present study are available upon reasonable request to the authors

